# Evidence-based Practice in Medical Education: Team-based Learning in a Blended Curriculum

**DOI:** 10.1101/2024.06.25.24309485

**Authors:** Natalie Smith

## Abstract

This article explores the redesign of a Physician Associate (PA) program’s didactic coursework from traditional lecture-based methods to a student-centered blended learning platform incorporating Team-Based Learning (TBL). The initiative, supported by a course innovation grant, aimed to address the limitations of passive learning by integrating active learning experiences that enhance student engagement and comprehension. The redesigned Diagnostic Methods I course leverages blended learning to combine online materials with face-to-face interactions, fostering critical thinking and practical application of knowledge. TBL, a structured approach to collaborative learning, facilitates deeper understanding through readiness assurance tests and team-based problem-solving. Early findings indicate improved student outcomes and positive perceptions of the blended TBL approach, suggesting its potential as a valuable educational strategy in PA programs. The article provides practical insights and lessons learned for educators seeking to implement TBL and blended learning in similar educational settings.

## INTRODUCTION

As clinicians, evidence-based medicine and science inform our clinical practice; likewise, as educators’ educational theory and learning philosophy should inform our teaching practice.^1^ The didactic phase of most Physician Assistant (PA) program curriculum still relies heavily on traditional lecture, comprised primarily of PowerPoint presentations and mandatory student attendance. High stakes student assessment in this context generally occurs in the form of exams after this passive learning format has taken place. This largely occurs without sufficient opportunity for student practice and application of the material through active learning experiences and without consistent instructor coaching or constructive feedback essential for deeper student understanding. Adult learning theory dictates that this approach to medical education is both antiquated and ineffective andragogy for PA educators.^1^

The author of this article was the recipient of a course innovation grant which provided the opportunity to redesign PA didactic coursework previously taught through traditional lecture-based methods. It is important to note that the support of the grant provided more time for the course director to initiate change but that the course innovation methods used through this process did not require additional personnel or technology aside from what was already being utilized by the program prior to acquisition of the grant. The changes were made by one PA faculty and are easily adaptable to other programs with limited resources.

This article reviews tips and key lessons learned through the process of course redesign from a traditional lecture course design to a student-centered blended learning platform featuring team-based learning (TBL). Practical improvement and course innovation are explored through the lens of what it is like to work within the confines of a busy PA didactic curriculum. TBL and blended learning are two high impact and evidence-based educational strategies that have demonstrated improved student outcomes in other facets of medical education but that are currently underutilized in PA education.^2^ After reading this article the PA educator will understand a practical approach toward effective utilization of TBL and blended learning as it applies to and improves student learning outcomes in PA medical education.

### The Course Innovation Process

The course that underwent re-design was Diagnostic Methods I which is a course designed to support the Clinical Medicine curriculum and focuses on appropriate ordering and interpretation of diagnostic studies. The course map below (**Figure 1**) demonstrates changes made based during re-design from a traditional lecture-based course.

**Figure 1.**
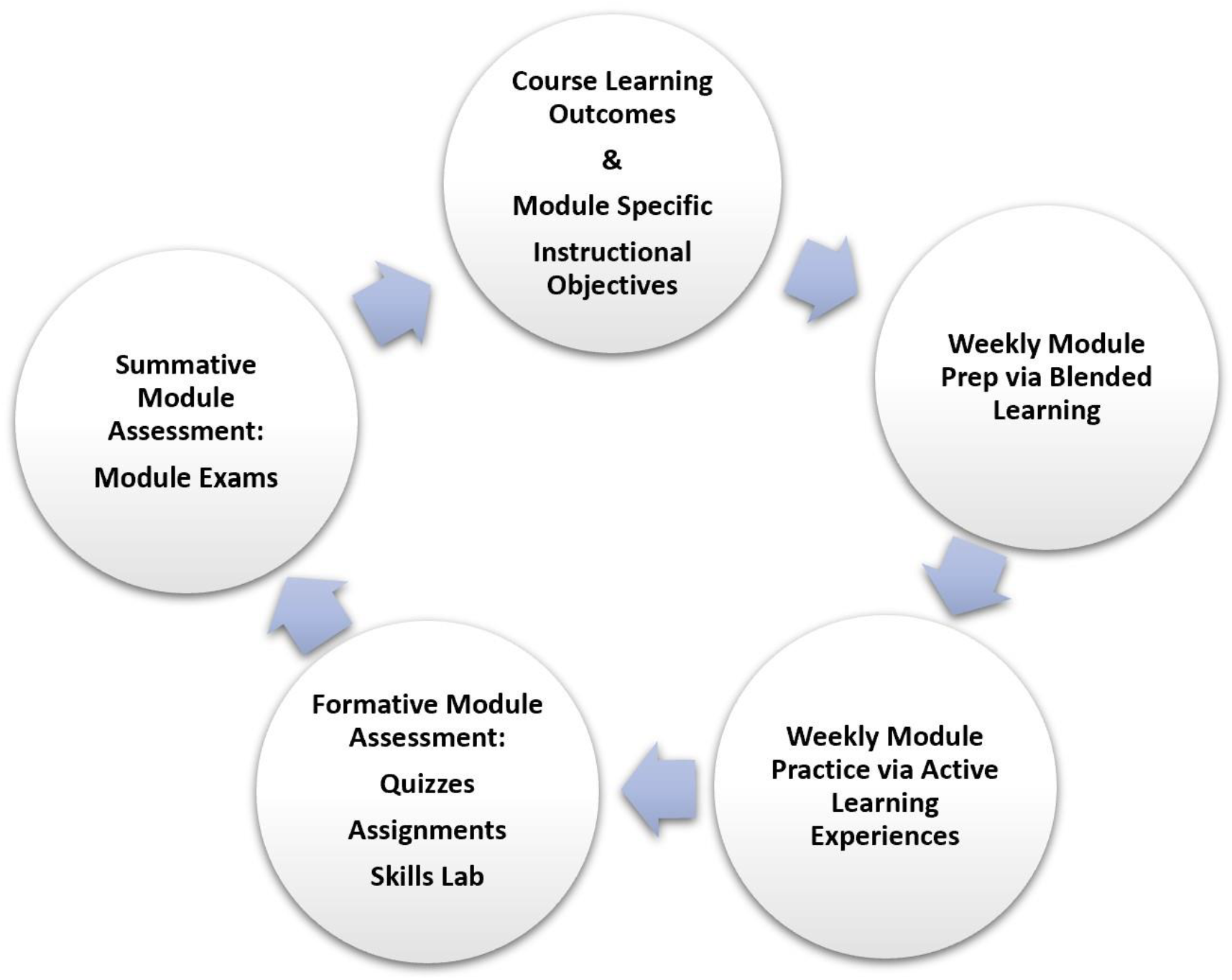
Diagnostic Methods I, course map after re-design.

### Blended Learning

Blended learning involves the integration of online materials and activities that are reinforced by frequent in person face-to-face classroom experiences. ^3^ This differs from hybrid learning where the online content is intended to replace large segments of face-to-face instruction, with students in person only for summative events such as assessments or graded OSCE’s. Blended learning in the PA curriculum is not only effective but has been shown to outperform traditional in-person learning practices in PA education.^4^ This approach allows students to maximize the benefits of online and face-to-face learning in a targeted way; a process which fosters the development of critical life-long learning skills. \

Posted online content should be diverse but not overwhelming and may take the form of short videos, podcasts, journal articles, assigned readings or mini lectures. Gamification and targeted use of online platforms and technology is a helpful adjunct to material presented as part of the blended learning process and serves as an important pillar to success with this learning format by facilitating student practice.

Practice experiences with the material also facilitate vital instructor coaching and constructive feedback. The author has used various modalities to assist with the blended learning strategy. Some of the most helpful have been: Anchor app for podcast creation through Spotify, Gimkit gaming for ECG practice, Flipgrid for acting out patient case presentations and student clinical skills recording, creation of virtual escape rooms (a student favorite), Quizizz and Kahoot! for asynchronous quiz items built into virtual mini lectures, etc. The more you can diversify the way you are delivering blended content, the more successful and engaging it will be.

### TBL: What is it?

TBL is uniquely equipped to allow for the integration of blended learning in a structured format.^4^ Once actually in the classroom, the students assimilate information in a social context with their peers and with a clinician educator being present for context, guidance, and debriefing.^5^ TBL offers PA educators an innovative teaching strategy that embodies the science behind what we know about effective learning and knowledge retention with consideration for both time and faculty resources.^6^

TBL can be effectively implemented in large classrooms, requiring only one faculty member to be present, with students divided into smaller teams (I typically run teams of around 7 students each). Students utilize modern learning practices based on constructivists learning theory by working in teams to process and apply their learning through the analysis of authentic clinical problems. This process requires negotiation, communication and achievement of team consensus within a short period of time.^2,7^

A distinguishing feature of TBL is the Readiness Assurance Testing or RAT. This process traditionally involves both an individual readiness assurance test (iRAT) and a team readiness assurance test (tRAT). The same test is given in each instance and is typically a short ten question quiz to assess preparation and to serve as low-stakes student practice prior to larger stakes examinations.^8^ **Table 1** provides a sample schedule for a traditional TBL format.

**Table 1.** Sample traditional 2-hour TBL schedule Online pre-class preparation materials reviewed by students.

|  |  |
| --- | --- |
| **10 min** | Individual readiness assurance test (iRAT) |
| **20 min** | Team readiness assurance test (tRAT) |
| **20 min** | Immediate review of iRAT/tRAT items |
| **60 min** | Active learning through authentic clinical problem-solving |
| **<u>10 min</u>**<br>**120 min** | Debriefing |

### Why use TBL?

TBL is a student-centered and evidence-based approach to medical education that fosters engagement and increases retention of knowledge.^9^ Improved student learning outcomes have been repeatedly demonstrated throughout the medical education literature with the use of TBL. Studies of medical school students have demonstrated not only that all medical students who learn through TBL achieve higher academic performance on examinations but that the performance benefit is maximized in students who are academically-at-risk and within the lowest academic quartiles.^2^ These students are forced to study more consistently and are provided with regular instructor coaching and performance feedback through integrated TBL practice. This provides systematic opportunity for growth and development of those coming into the TBL experience with less content mastery.^2,9^ These students also benefit the most from the built in peer-to-peer teaching that occurs through collaboration, debate and team decision making with their peers who are already achieving academic success.^2^

### How is TBL different?

TBL has been shown to outperform its more commonly recognized counterparts, case-based learning (CBL), problem-based learning (PBL) and traditional lecture.^10,11^ TBL more than CBL or PBL places emphasis on assuring students have a solid understanding of the material through the use of integrated practice and readiness assurance testing with immediate feedback and review.^10^ Additionally iRAT testing has been shown to be a reliable predictor of later exam performance and serving as valuable practice for the student and also as a critical check-point enabling self-assessment with time for correction and instructor coaching prior to the examination.^9^ TBL improves student understanding of difficult concepts and improves examination performance on concepts aligned with TBL activities.^2,10–12^ TBL has also demonstrated consistency in positive student perceptions of instruction, with students reporting the events to be more rewarding in terms of a deeper understanding of concepts, more motivating in terms of a sense of responsibility to their team, and more interactive than regular case-based discussions (CBL) or PBL.^8,11^

### How should I Incorporate TBL as a PA educator?

There are many nuances to PA education that can make a modified version of TBL more effective for our students. The modified TBL format for PA educators proposed below (**Figure 2**) reflects the experience of the author with the traditional TBL format and observations made which are specific to PA learners and our curriculum pace. Changes to the traditional format (**Table 1**) were necessary to create a model reflective of the challenges unique to PA education.

**Figure 2.**
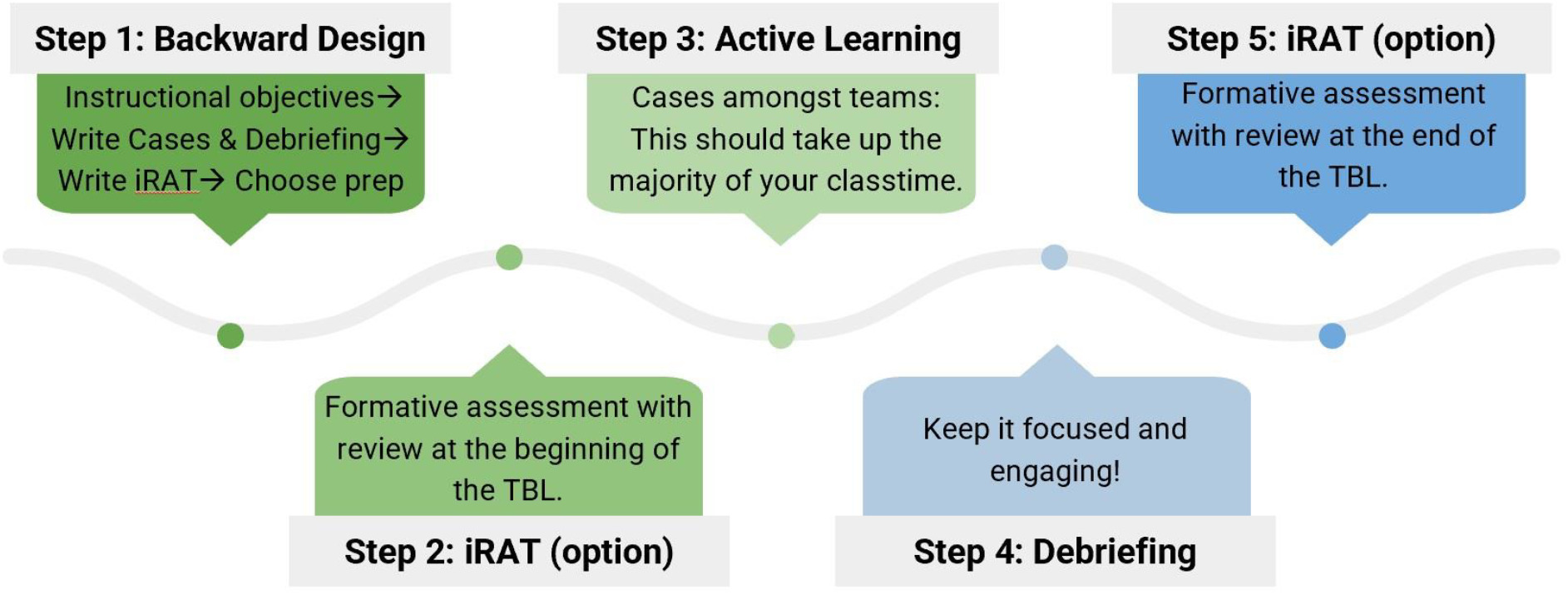
Modified TBL format for PA educators

An effective TBL lives and dies by effectively conveyed instructional objectives (see **Table 2**). Choose relevant module instructional objectives and post those for student TBL preparation and guidance. Do not be afraid to “give away” the cases of the session with objectives that are seemingly too specific. Remember, the point is for the students to practice and to learn the material, there should not be surprise topics on the quiz or with case diagnoses-it is the practice of getting to the diagnosis that is important. To be effective, TBL instructional objectives need to be focused and clear. To do this the objectives need to be concise and clinically relevant (what do they *really* need to know to be an effective entry level PA?) Objectives should be posted well in advance of the event so that students know exactly how to best prepare for the session.

**Table 2.** Sample TBL Instructional Objectives, Module 3: Cardiovascular.

| **Module Instructional Objectives (MIO)** |  |
| --- | --- |
| **MIO3.4** | Evaluate the clinical indications, clinical utility and interpretation of diagnostic findings related to congestive heart failure (imaging and lab diagnostics). |
| **MIO3.5** | Formulate an order set for work-up of vascular disease presentations (arterial vs. venous) and be able to differentiate between signs and symptoms of the two. |
| **MIO3.6** | Evaluate the clinical indications for the chest-pain work-up. |
| **MIO3.7** | Formulate an inclusive order set based upon your appropriate differential diagnosis of chest pain and interpret the resultant findings. |

It is also important to use backward design when planning a session. The educator should choose session instructional objectives first and then develop the case content in its entirety (not just the diagnosis). Develop the important take-home points of the debriefing and finally then write the quiz questions focused on the need-to-know concepts for solving the activity or case.

When writing the quiz, avoid picky questions that focus only on briefly mentioned content in the preparation materials that were not reinforced within the cases or the debriefing. Only after writing the quiz, should you then choose and assign the relevant preparation content. This order is surprisingly important. The author has done this incorrectly in the past by first assigning the students preparation material (based on case diagnoses I had in mind), and then writing the quiz questions based on the preparation material prior to developing the actual in-depth case content (beyond diagnosis) for the session. This mistake led to an unintentional tendency to ask less “big picture” questions on the quiz. The quiz was then perceived by some students as difficult and inconsistent with what they had perceived to be important with preparation work which resulted in a defeated tone for those students at the very beginning of the session, even before the active learning TBL session had a chance to engage their minds further.

One of the aspects of PA education that becomes difficult within a traditional TBL framework is our expectation for student proficiency and assimilation of large volumes of material at a pace that exceeds that of many other professional program curricula. TBL in the traditional format (see **Table 1**) utilizes both the iRAT and tRAT as a means of formative assessment and collaborative decision making.^3^ Although this process did generate in depth conversation and debate amongst teams regarding the quiz items, there was an inordinate amount of time spent on the 10-question quiz and the process of taking it twice. In the author’s experience, this approach distracted from the instructional objectives of the session and placed unintended emphasis on the quiz and its associated grade for students. The author has found that by omitting the tRAT and administering the iRAT only there is ample assurance that pre-class preparation has occurred, and the benefit of increased time spent on the active learning components.

Moving the timing of the iRAT to the end of the session is another modification that the author has seen foster engagement. This change promotes buy-in from students. It reinforces the idea that the focus is not only to prepare for a quiz grade (extrinsic motivation for learning) but to practice and experience meaningful application and assimilation of the material through the active learning process (resulting in a shift toward intrinsic motivations for learning). In the authors experience, students come to class just as prepared as there is still an individual quiz grade and the element of team accountability throughout the session. Additionally, there is enhanced engagement during the cases and the debriefing because the students do not want to miss any teaching points that may be reiterated on the iRAT. By moving the iRAT to the end of the session I find students asking more key foundational questions and asking the instructor to repeat things again to make sure they have the proper understanding prior to the quiz. They end up wanting the same thing I do, to maximize their learning. **Figure 3** from student survey data shows student perceptions of this TBL modification. There were 33 students in the cohort that experienced the curriculum change and were surveyed. The survey response rate was 30 students.

**Figure 3.**
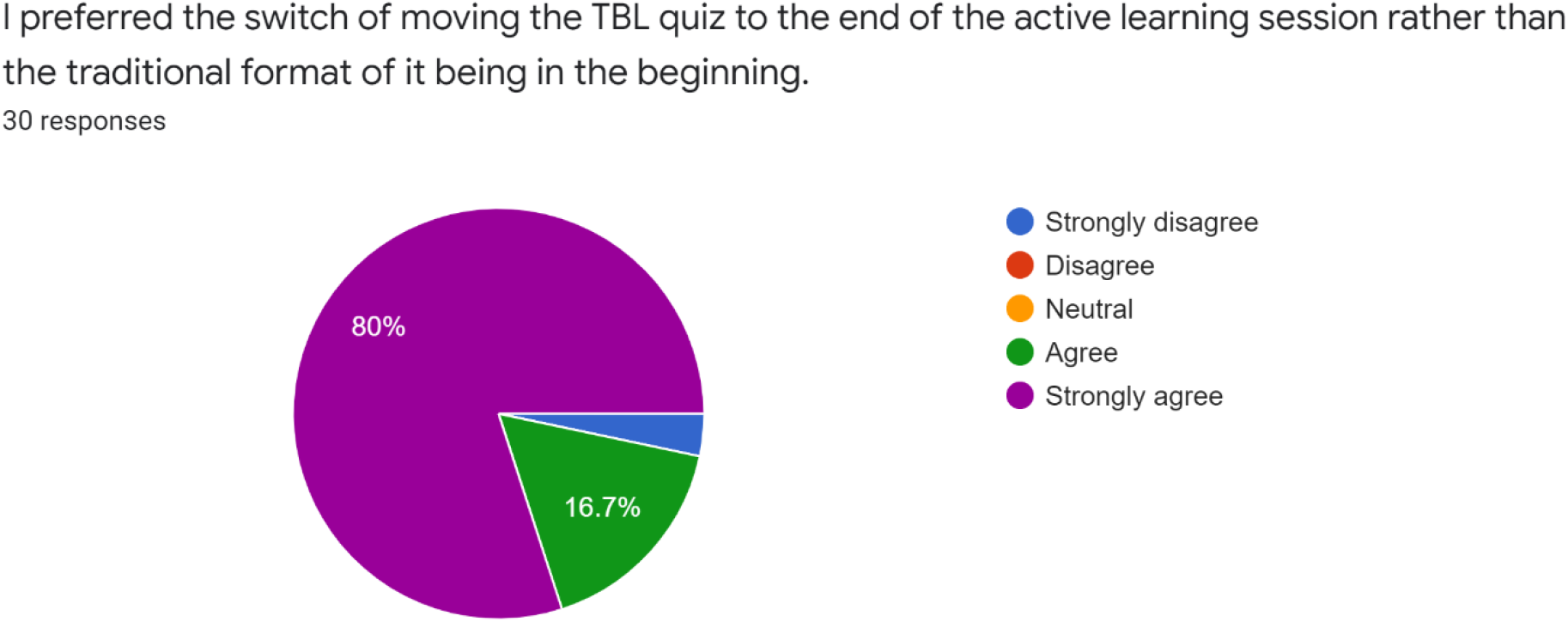
Student perception, iRAT modification.

**Figure 4.**
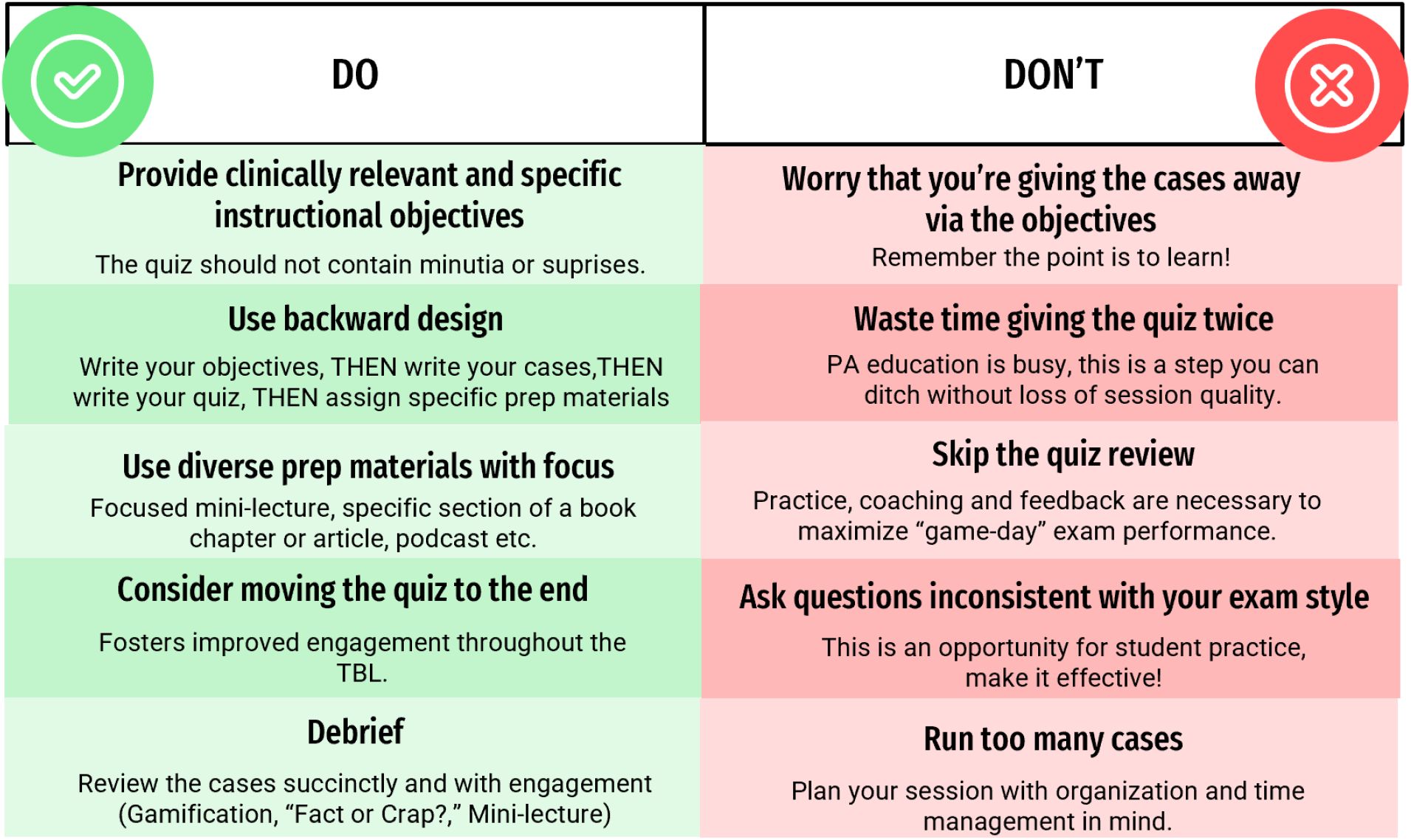
Dos and Don’ts of TBL for the PA Educator

The debriefing should be a succinct summary of the major teaching points of the cases and instructional objectives. This may take the form of a mini-lecture or an interactive game or exercise for wrap-up. One simple example I created is a game of “Fact or Crap” where the students are given corresponding signs made by taping a “Fact” or “Crap” emoji to tongue depressors. The students are then presented with a series of statements that summarize the major teaching points of the session and hold up their respective signs to indicate a more engaging version of “true or false”. Another way to get them up and moving for the debrief is to have everyone stand up, present a series of statements, and have the students respond as true or false by remaining standing or by sitting down. These are opportunities to give feedback and make clarifications. Constructive feedback should be used as a tool throughout the sessions for improved team performance and deeper student understanding.

### Early Findings

Student perceptions of the curriculum change to TBL in a blended learning format were evaluated by student survey. Survey analysis results are depicted below. There were 33 students in the cohort that experienced the curriculum change. The survey response rate was 30 students. Overwhelmingly students felt engaged with TBL and felt it added value to their learning and understanding of the material (**Figure 5)**. Furthermore, they recommended it’s use as a teaching modality in the future (90%, **Figure 6**).

**Figure 5.**
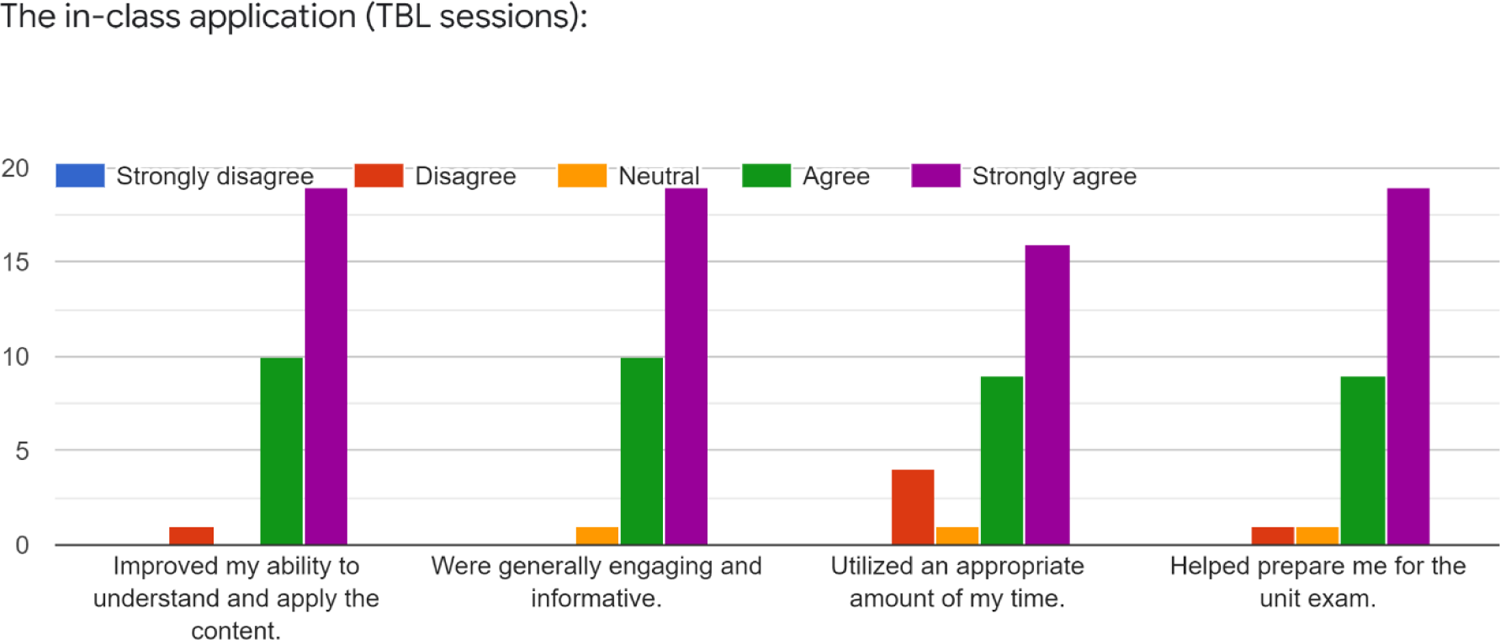
Student perception of TBL

**Figure 6.**
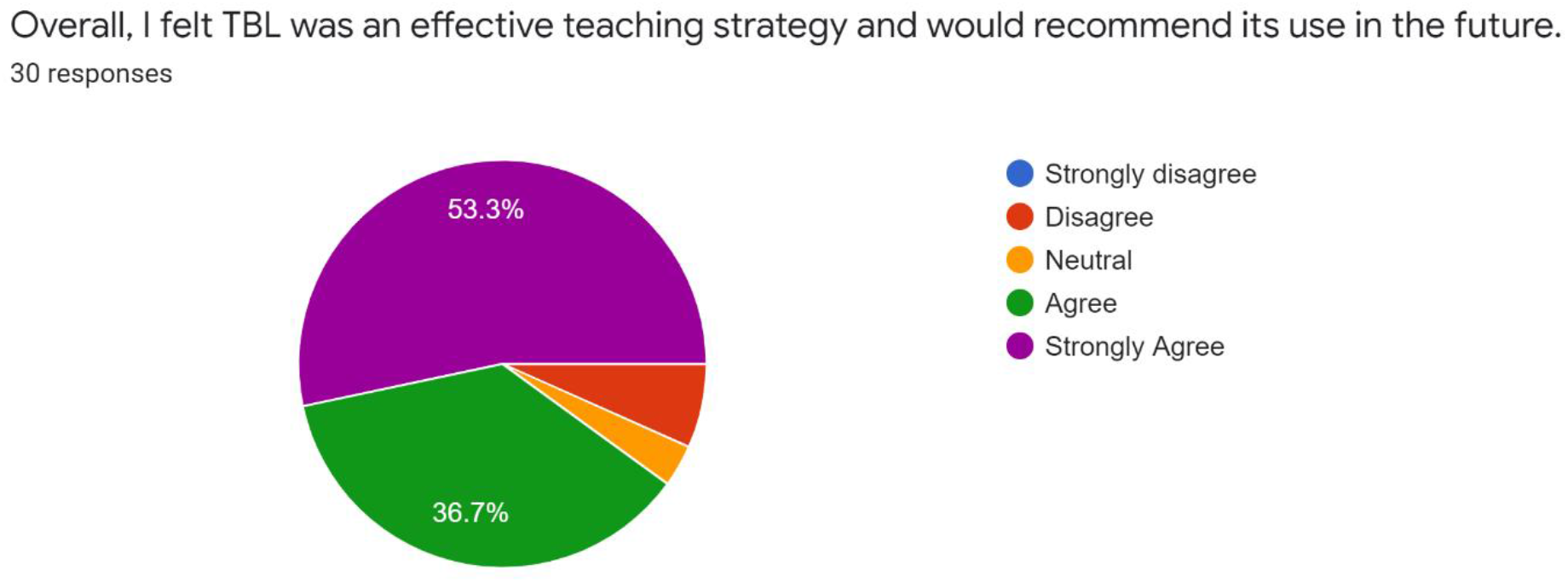
Student opinion, was TBL effective?

The early findings related to this curriculum change also show improved PA student outcomes. Exam scores were compared to previous cohorts learning the same material with a traditional lecture-based format and with the same instructor. Findings related to modular exam outcomes are summarized in **Table 3** below.

**Table 3.** Exam Averages, Traditional Instruction vs. TBL and Blended Learning.

|  | 2019 | 2020 | 2021 | TBL 2022 |
| --- | --- | --- | --- | --- |
| **Module 1 Exam Avg.** | 90% | 87% | 89% | 92% |
| **Module 2 Exam Avg.** | 88% | 87% | 87% | 89% |
| **Module 3 Exam Avg.** | 91% | 90% | 93% | 92% |
| **Final Exam Avg.** | 89% | 91% | 89% | 92% |

**Table 4.** Lowest Exam Performance, Traditional Instruction vs. TBL and Blended Learning.

|  | **2019** | **2020** | **2021** | **TBL 2022** |
| --- | --- | --- | --- | --- |
| **Module 1 Exam Low Score** | 76% | 71% | 82% | 85% |
| **Module 2 Exam Low Score** | 79% | 68% | 76% | 75% |
| **Module 3 Exam Low Score** | 82% | 72% | 82% | 84% |
| **Final Exam Low Score** | 81% | 79% | 77% | 83% |

Exam 1, 2, 3 and final exam averages across the previous cohorts (2019, 2020, and 2022) were: 88%, 87%, 91%, and 90% respectively. The year 2022 (TBL) shows a 4% increase from the exam 1 average (2019-2021), 2% increase in the exam 2 average (2019-2021), 5% increase in exam 3 (2019-2021), and a 2% on the final exam (2019-2021).

It is also interesting to reflect on how the curriculum redesign may have affected those students in the lower quartiles of performance. The averages of the lowest exam scores across previous cohorts (2019, 2020, and 2022) were: 76.3%, 74.3%, 78.6%, and 79.0% respectively. The lowest performances in the year 2022 reflect: a 9%, 8%, 7%, and 4% increase of lowest exam averages of these previous cohorts on exams 1, 2, 3, and the final exam respectively.

The limitations of this early assessment include the need for more robust, formal statistical analysis currently limited by incomplete available data at the time of the article as well as assessment of any differences in cohort profiles.

## CONCLUSION

Active, experiential learning practices are necessary to harness student engagement and produce a higher level of understanding and applied critical thinking. PA educators need to recognize these evidence-based findings as opportunities to embrace a more student-centered and effective andragogy. TBL demonstrates improved student outcomes compared with CBL, PBL and traditional lecture environments in PA education by utilizing the science of adult learning theory.^4^ A modified TBL framework should be integrated within PA education coursework as a practical component of a larger strategy involving blended learning, opportunities for student practice and formative assessment as well as ongoing instructor coaching and constructive feedback.

## Data Availability

All data produced in the present work are contained in the manuscript.

